# The Art of Pain: A Quantitative Colour Analysis of the Self-Portraits of Frida Kahlo

**DOI:** 10.1101/2022.07.21.22277897

**Authors:** Federico E. Turkheimer, Jingyi Liu, Erik D. Fagerholm, Paola Dazzan, Marco L. Loggia, Eric Bettelheim

## Abstract

Frida Kahlo (1907-1954) was a Mexican artist who is remembered for her self-portraits, pain and passion, and bold, vibrant colours. This work aims to use her life story and her artistic production in a longitudinal study to examine with quantitative tools the effects of physical and emotional pain (rage) on artistic expression.

Kahlo suffered from polio as a child, was involved in a bus accident as a teenager where she suffered multiple fractures of her spine and had 30 operations throughout her lifetime. She also had a tempestuous relationship with her painter husband, Diego Rivera. Her physical and personal troubles however became the texture of her vivid visual vocabulary — usually expressed through the depiction of Mexican and indigenous culture or the female experience and form.

We applied colour analysis to a series of Frida’s self-portraits and revealed a very strong association of physical pain and emotional rage with low wavelength colours (red and yellow), indicating that the expression of her ailments was, consciously or not, achieved by increasing the perceived luminance of the canvas. Further quantitative analysis that used the fractal dimension identified “The broken column” as the portrait with higher compositional complexity, which matches previous critical acclaim of this portrait as the climax of her art. These results confirm the ability of colour analysis to extract emotional and cognitive features from artistic work. We suggest that these tools could be used as markers to support artistic and creative interventions in mental health.

## INTRODUCTION

### PREAMBLE

Creating visual art is one of the defining characteristics of the human species [18] and is layered by both conscious and unconscious activities [24], so that canvases may contain elements stemming from the physical and mental ailments of the artist. We now have the tools to extract and resolve these by use of quantitative tools [27].

From the early decoration of human bodies with skin colouring and beads and the early 2-dimensional forms created at least 30,000 years ago to the increasing sophistication of images represented on various surfaces and materials, visual art has been the vehicle of expression for the historical and social narratives of civilizations, their aesthetics, and their values{Sigaki, 2018 #90;Kim, 2014 #89;Morriss-Kay, 2010 #84}. At the same time, artistic production may be the vehicle for the author’s personal and intimate narratives and mirror her/his craft and unique creative ambitions, as well as inner impulses and subconscious mental states — represented by the Art Brut collection in Lausanne [6], that contains multiple art pieces created by individuals with psychiatric disorders.

We suggest that it is of interest to develop quantitative approaches that can extract from visual art features strongly associated with underlying pathology that, besides the inherent cultural interest, may ultimately inform the use of art as a therapeutic intervention in mental health [31].

### ART, NEUROPATHOLOGY AND THE LONGITUDINAL DESIGN

Features extracted from works of art can be simply associated to certain diagnostic categories {Hacking, 1996 #124}, but they become true biomarkers when they can be meaningfully connected to underlying pathology. For example, in the case of the dementias, Gretton and fytche [11] were able to distinguish changes in the use of the canvas, the colour palette and artistic themes amongst artists with Alzheimer’s disease, frontotemporal dementias and Lewy Bodies disease and link these features to neurological deficits and imaging findings. This insight is harder to achieve in psychiatric conditions as pictorial characteristics are not necessarily closely associated with diagnosis [32]; this likely stems from the heterogeneous and dynamic nature of these disorders and the far less defined neuropathology [1; 29]. However, it turns out that pictorial features are of great value in longitudinal studies that provide a dynamic understanding of the patient, regardless of diagnosis [32]. In a seminal longitudinal study of ∼200 patients with unipolar depression, bipolar depression and acute schizophrenia, conducted at N.I.H. in the 70’s, Wadeson and Carpenter used the qualitative categories of colour, fullness of space utilized, extent of development and organization to score >1,000 pieces produced by the participants [32].

They were not able to use these pictorial characteristics to reliably classify participants in the three diagnostic groups but the longitudinal information extracted for each participant was found to be helpful in the planning of their treatment; in their own words:

> “*the art evaluations have proved most useful in comparing pictures made by the same patient at different times thereby eliminating variables of artistic ability, art experience, hand-eye coordination, intelligence and other factors that may affect comparisons across individuals”* [32].

### QUANTITATIVE APPROACHES FOR THE ANALYSIS OF LIFE-TIME ARTISTIC TRAJECTORIES

It is clear that art can be effectively integrated into psychiatric lines of inquiry if a) experimental designs focus on the individual trajectory of the author’s compositions and her/his symptoms, b) the methods used are quantitative as variation of features is likely to be subtle and c) features can be linked to the underlying neurobiology.

The value of life trajectories in artistic analysis is now well recognized [14]. Computational methods that extract quantitative features from canvases are commonly used by art collectors, art dealers and museums for the detection of forgeries but also in art history to identify styles and artists, determine cross-influences, and, more experimentally, have been applied to predict aesthetic ratings (please see review of these methods in [2]). Recently, we have been able to integrate all three steps above in a longitudinal, quantitative study of the self-portraits of Vincent van Gogh to demonstrate his mental illness as likely due to interneuron deficits due the significant association of colour contrast and expressionist style with his consumption of absinthe, a strong liquor with potent activity on gamma-aminobutyric acid (GABA) receptors [27].

### FRIDA KAHLO AND THE ART OF PAIN AND ANGER

Here we present a life-time analysis of the art of the Mexican painter Frida Kahlo (1907-1954) that utilizes quantitative tools to investigate the relationship between the colour palette and the artist’s mental health at various times of her life.

As before, we have chosen to restrict the analysis to the very rich collection of self-portraits that the artist produced to exert some control over the subject matter of the art. We then projected digital copies of these portraits in the three cardinal colour spaces and quantified features. These features were used in a fashion different from the previous work on van Gogh as here the question was not about neuropathology. We have instead used Frida Kahlo’s medical history to establish how physical pain and anger may have percolated into her painting style and used detailed biographical notes to establish those colour features best associated with her medical and life history.

### HYPOTHESIS

The hypothesis tested here was that the colours with high perceptual intensity, and red in particular, would be present in the self-portraits of artist Frida Kahlo as the result of the painter’s physical pain or deep emotional anger.

Colours are an ubiquitous part of our percepts and it has been known for some time that they have differing effects on human cognition and behaviour (please see [8] for a review). Colour also has a modulating effect on pain and, depending on wavelengths, visual stimuli may induce analgesia or hyperalgesia [5]. In particular, visual application of red light (long wavelength at 660 nm) has been shown to exacerbate pain in both human and animal models, and even induce functional pain in an injury-free model [13; 33] through a number of pathways that modulate pain processing in the cerebrum and brainstem and reflect on parasympathetic and sympathetic functions with the release of a number of neurotransmitters and hormones (e.g. dopamine, GABA, histamine, orexin, melanin-concentrating hormone, oxytocin, and vasopressin) [16; 20-22].

Interestingly, the colour red also seems to exert effects at higher levels of cortical processing and modulate the response to affective images [8; 9]. There is no agreed mode of explanation for this association as this colour could appear in emotional situations with evolutionary significance (e.g. red blood) or may become arbitrarily associated in individual cultures (e.g. red faces in angry people of Caucasian tribes), or both [12], ultimately becoming part of a human process of grounding abstract concepts (e.g. moods) in perceptual experience [17]. The exact mechanisms of regulation of mood by light perception and vice versa have not been precisely worked out, but there is ample evidence that they impinge on the same sympathetic and parasympathetic pathways involved in pain processing [7].

## METHODS

To test the hypothesis, two parallel data sets were employed, a series of colour self-portraits together with biographical material and detailed catalogue notes. The former dataset was treated with standard computational tools to extract colour features while the information contained in the latter was used to score (0 – no pain, anger, 1 – yes pain and anger) to determine whether the physical and/or emotional pain was expressed intentionally or sub-consciously in the individual art piece.

### LIFETIME TRAJECTORY AND MEDICAL HISTORY

Few artists of the 20^th^ century have had their personal and private lives as closely examined as the Mexican painter Frida Kahlo (1907-1954). Her career was relatively short with an output of little more than 150 paintings; of these 55 were self-portraits. Yet her aesthetic and conceptual ideas have inspired an ever-growing fascination across the world as her career was marked by a series of upheavals that were intimately connected to her private life, hence the intense scrutiny and interest concerning even the most minute biographical detail. Here we will focus on three main factors that dominated her physical and mental health during her life: pain, heartache and anger as described in the detailed clinical analysis written by V. Budris [3].

Frida Kahlo was born in 1907 with a congenital anomaly, spina bifida that manifests with several neurological and skeletal abnormalities, in her case in the lower part of the body and feet. Aged six, she then caught poliomyelitis that aggravated the deformity on her right leg and occasionally caused discomfort and pain. Later in her life, she underwent several surgical procedures on her right foot and leg that eventually led to gangrene and amputation of her leg shortly before her death in 1954.

In 1923 at the age of 18, she was involved in a bus accident where she suffered severe injuries to the spine and further damage to her right leg. The forced confinements in bed during the following months made her change her career path, abandoning medical school and turning to painting as a way to deal with boredom and pain. Later she joined the local Communist Party where she met Diego Rivera, an established Mexican artist 30 years her senior, who become her husband in 1929, and the love of her life. The state of her spine and pelvis deteriorated throughout the years, was the source of excruciating pain at times and caused three miscarriages ultimately preventing her from having any children. Between 1946 and 1951 she underwent eight surgical procedures that were meant to ameliorate her deteriorating condition but, ultimately, made her situation worse as she became confined to a wheelchair.

The meeting with Diego Rivera had a major professional impact as his international reputation and his contacts with American and European circles facilitated the propulsion of Frida into fame. However, the relationship was tumultuous with Diego’s multiple extra-marital affairs; one in particular with Frida’s sister Cristina, once her closest confidant, which enraged Frida and caused their divorce in 1939 only for them remarry again one year later. However, the events leading to the separation had changed Frida. She was now far more independent, both personally and professionally, carrying on several romantic liaisons of her own, one famously with Leon Trotsky.

Frida Kahlo was a key figure of Mexican revolutionary modern art and international surrealism, her canvases expressing her personal turmoil but also mirroring her philosophy and vision as she was a pioneer of the politics of gender, sexuality, and feminism. To isolate/identify objectively the expression in her art of the physical and emotional factors of interest, we relied on the most complete catalogue of her production recently published by Taschen [15]. In the catalogue, we searched the extensive notes that reference each self-portrait and scored the art piece with a pain factor of 1 if the notes clearly referred to an intent to express pain, heartache, or anger.

### COLOUR EXTRACTION AND FEATURES

The hypothesis to be tested was about a potential association of colours of high intensity and the painter’s physical pain or deep emotional anger.

This required the extraction of features related to the colour red. The colour itself can be characterized by hue (wavelength) and redness (contribution of the primary colour red); moreover, the effect of the colour could be exerted by its overall perceptual lightness or by the light contrasts created by the painter in the canvas.

Forty-three self-portraits that are in colour, on canvas and catalogued [15] were downloaded from the on-line digital exhibition of her work available at www.FridaKahlo.org.

The digital self-portraits were then analyzed to extract the relevant features following established protocols [23]. The paintings were first analyzed the original RGB space, in which the contents of the Red, Green and Blue (RGB) channels were separated from one another and mean values for each channel calculated.

The data were then projected into the Hue, Saturation, Value (HSV) space, in which we calculated the percentage of voxels contained in the first bin of the hue channel to estimate the number of red pixels in the painting [23].

The data were then projected into the CIELAB space that also has three dimensions called L*, a* and b*. The perceptual lightness value, L*, defines black at 0 and white at 100, the a* axis defines the green–red spectrum, and the b* axis represents the blue–yellow spectrum. Here we calculated the mean perceptual lightness of L*.

To quantify the contrast of the luminance in the portraits, we calculated the average contrast across the image. For each digital image, the luminance of a pixel was obtained from its red, green, blue components as 0.299R + 0.587G + 0.114B (International Telecommunication Union, Recommendation BT .601). We then calculated the power spectrum of the image intensity using the 2-D Fourier Fast Transform (FFT) and the value at the origin of the FFT produced the required average image contrast [4; 27].

Furthermore, we examined the canvases for indications of neurological deterioration following the worsening of her condition, particularly after 1945 and the numerous and ineffective surgical procedures during the last years before her death [3]. To this end, we calculated the fractal dimension (FD) of each self-portrait using a previously described procedure specific to RGB images [19]. Fractals are self-repeating patterns across scales that are the hallmark of certain natural objects or organisms such as snowflakes, river beds, and coastal lines, as well as the human brain and its functional signatures, from neuronal activity to motor behaviour, such as writing and painting [10; 26; 28; 29]. The FD quantifies the space filling property of the object measured; from a neurological perspective, larger FD indicates the ability of the artist to control the object on canvas in a cohesive fashion and meaningfully connect all the scales of its realization from small details, to the large scenery or background [10]. Longitudinal analysis of the FD has revealed either a stable or rising trajectory in the work of painters with no obvious medical history, the latter likely indicating increasing artistic maturity, while decreasing values were found in those with neurological or neurodegenerative conditions [10].

### ANALYSIS

The process described above produced the following 7 features: red-channel, green-channel, blue channel mean intensities, luminance contrast, mean perceptual lightness, percentage of red pixels and fractal dimension.

These features were then tested for differences between the two groups of portraits, one expressing pain/anger (pain score = 1) versus the control group (pain score =0). Given the normal-like distribution of the scores, independent group Student t-tests were conducted, preceded by a Levene test for equality of variances. In the case of FD, the effect of time (in units of years) was controlled for prior to testing using a standard Pearson correlation test. Features were considered significant if the resulting *p*-value was <0.05/*n* where *n* = 7 accounts for the increased expected number of false positives given the multiple tests and implements the Bonferroni correction [30]. All calculations were performed using Matlab (v. R2018b, The Mathworks Inc., Natick, MA, USA) while statistical analyses were conducted in SPSS v.25 (IBM Corp).

## RESULTS

A total of 43 self-portraits were identified, downloaded, and processed. Table 1 contains the list of the portraits together with the corresponding year of production, pain/rage score, the catalogue references where text clearly referred to pain or anger and a brief note of explanation. Out of these, about half (*n*=22) had a positive score as the catalogue entries clearly referred to the intent of Frida to express pain or anger [15]. This could have been for a number of reasons including the anger about a miscarriage or the physical pain accompanying the many surgical procedures she endured, or the anger towards her unfaithful husband.

**Table 1:** The list of the self-portraits with corresponding year of production, pain/anger scores that were assigned based on the intent of the author to express pain or anger, the reference the scores were based upon (pages and catalogue numbers as in [15]) and brief notes on what the pain score is based upon.

| **Title** | **Year** | **Score** | **Reference** | **Notes** |
| --- | --- | --- | --- | --- |
| Self portrait in a velvet dress | 1926 | 0 | pp471, cat.4 | - |
| Self portrait time flies | 1929 | 0 | pp.491, cat.28 | - |
| Frieda and Diego Rivera | 1931 | 0 | pp.494, cat.32 | - |
| Self portrait along the border line between Mexico and the United States | 1932 | 0 | pp.500, cat.37 | - |
| Henry Ford Hospital | 1932 | 1 | pp.502, cat.39 | Miscarriage |
| Self-portrait with necklace | 1933 | 0 | pp.508, cat.43 |  |
| Self portrait with curly hair | 1935 | 1 | pp.511, cat.46 | Anger for treason |
| A few small nips - passionately in love | 1934 | 1 | pp.511, cat.47 | Anger for treason |
| Fulang Chang and I | 1937 | 0 | pp.517, cat.52 | - |
| My nurse and I | 1937 | 0 | pp.520, cat.54 | - |
| Me and my doll | 1937 | 1 | pp.521, cat.55 | Miscarriage |
| Memory the heart | 1937 | 1 | pp.523, cat.56 | Anger for treason |
| Self-portrait dedicated to Leon Trotsky | 1937 | 0 | pp.524, cat.57 | - |
| Self-portrait the frame | 1938 | 0 | pp.535, cat.68 | - |
| Self-portrait with a monkey | 1938 | 0 | pp.537, cat.70 | - |
| Itzcuintli dog with me | 1938 | 1 | pp.536, cat.69 | Miscarriage |
| The two Fridas | 1939 | 1 | pp.546, cat.78 | Divorce |
| The dream the bed | 1940 | 1 | pp.549, cat.80 | Divorce |
| Self-portrait dedicated to dr Eloesser | 1940 | 1 | pp.550, cat.81 | Divorce |
| Self-portrait with necklace of thorns | 1940 | 1 | pp.551, cat.82 | Divorce |
| Self portrait with monkey | 1940 | 0 | pp.553, cat.83 | - |
| Self-portrait dedicated to Sigmund Firestone | 1940 | 1 | pp.554, cat.84 | Divorce |
| Self-portrait with cropped hair | 1940 | 1 | pp.555, cat.85 | Divorce |
| Self-portrait with braid | 1941 | 0 | pp.556, cat.86 | - |
| Me and my parrots | 1941 | 0 | pp.557, cat.87 | - |
| Self-portrait with Bonito | 1941 | 0 | pp.558, cat.88 | - |
| Self-portrait in red and gold dress | 1941 | 0 | pp.558, cat.89 | - |
| Self-portrait with monkey and parrot | 1942 | 0 | pp.563, cat.96 | - |
| Self-portrait with monkeys | 1943 | 0 | pp.564, cat.97 | - |
| Roots | 1943 | 1 | pp.566, cat.99 | Physical Pain |
| Self-portrait as a Tehuana | 1943 | 1 | pp.567, cat.101 | Divorce (portrait started in 1940) |
| Thinking about death | 1943 | 0 | pp.569, cat.102 | - |
| The broken column | 1944 | 1 | pp. 574, cat.110 | Physical Pain |
| Without hope | 1945 | 1 | pp.584, cat 120 | Physical Pain |
| Tree of hope remain strong | 1946 | 1 | pp.586, cat.121 | Physical Pain |
| The wounded deer | 1946 | 1 | pp.587, cat.122 | Physical Pain |
| Self-portrait with loose hair | 1947 | 0 | pp.589, cat.123 | - |
| Self-portrait in medallion | 1948 | 1 | pp.591, cat.125 | Physical Pain |
| Diego and I | 1949 | 0 | pp.594, cat.127 | - |
| The love embrace of the universe the earth Mexico myself Diego and- señor Xolotl | 1949 | 1 | pp.592, cat.126 | Physical Pain |
| Self-portrait with the portrait of doctor Farill | 1951 | 1 | pp.598, cat.131 | Physical Pain |
| Self-portrait with a portrait of Diego on the breast and Maria between the eyebrows | 1954 | 1 | pp.609, cat.145 | Anger |
| Self-portrait with Stalin | 1954 | 0 | pp.610, cat.148 | - |

The results of the feature comparison between the two groups of paintings are reported in Table 2 in terms of mean values ± standard deviations, student-t statistic and *p*-values. As expected, redness (mean of the R channel) was very much increased in the pain/anger group of portraits (Cohen’s d 1.33); however so was the green channel (Cohen’s d 1.27) and the blue channel (1.11). This confirmed the qualitative impression that in this group, Frida Kahlo made abundant use of yellow (R+G) and white (R+G+B). In fact, the difference between the two groups is better captured by the increase in perceived lightness (L*), that corresponds to whiteness. The red hue, e.g., the percentage of red pixels, was unchanged across groups. The fact that lightness and not hue varied between groups is further supported by a substantial increase of luminance contrast in the pain/anger portraits. In other words, at least for Frida Kahlo, the expression of physical and emotional pain was obtained by increasing the total perceived luminance and the localized luminance contrast on the canvas.

**Table 2:** Results of the group comparison of features between self-portraits scored 0 for pain and anger and those scored 1. Results are reported as mean values ± standard deviation, Student-t statistic and p-value. P-values smaller than 0.05/7 (0.007) are indicated with the double asterisk and are significant as they survive Bonferroni correction.

| Features | Score 0 | Score 1 | T ($d.f.=41$) | P-value |
| --- | --- | --- | --- | --- |
| **R-Channel** | 115.8 ($\pm 20.6$) | 147.4 ($\pm 26.3$) | 4.37 | 0.00008** |
| **G-Channel** | 94.2 ($\pm 24.1$) | 123.6 ($\pm 24.1$) | 4.20 | 0.00015** |
| **B-Channel** | 70.7 ($\pm 26.6$) | 100.5 ($\pm 26.8$) | 3.66 | 0.00071** |
| **Contrast** | 0.195 ($\pm 0.080$) | 0.303 ($\pm 0.092$) | 4.09 | 0.00017** |
| **Lightness** | 40.2 ($\pm 10.7$) | 55.3 ($\pm 11.7$) | 4.39 | 0.00007** |
| **Red Pixels (%)** | 0.47 ($\pm 0.18$) | 0.46 ($\pm 0.21$) | 0.09 | 0.92 |
| **FD** | 2.39 ($\pm 0.22$) | 2.43 ($\pm 0.18$) | 0.61 | 0.55 |

Figures 1 and 2 demonstrate the timeline for the self-portraits in relation to perceived luminance L* and R channel intensity. Figure 3 and Figure 4 display two portraits each, at the top and bottom of the luminance and the redness scales. The portraits were chosen as they display very similar themes, but a completely different mood is clearly reflected in the lightness that is high in the first set and much lower in the second.

**Figure 1:**
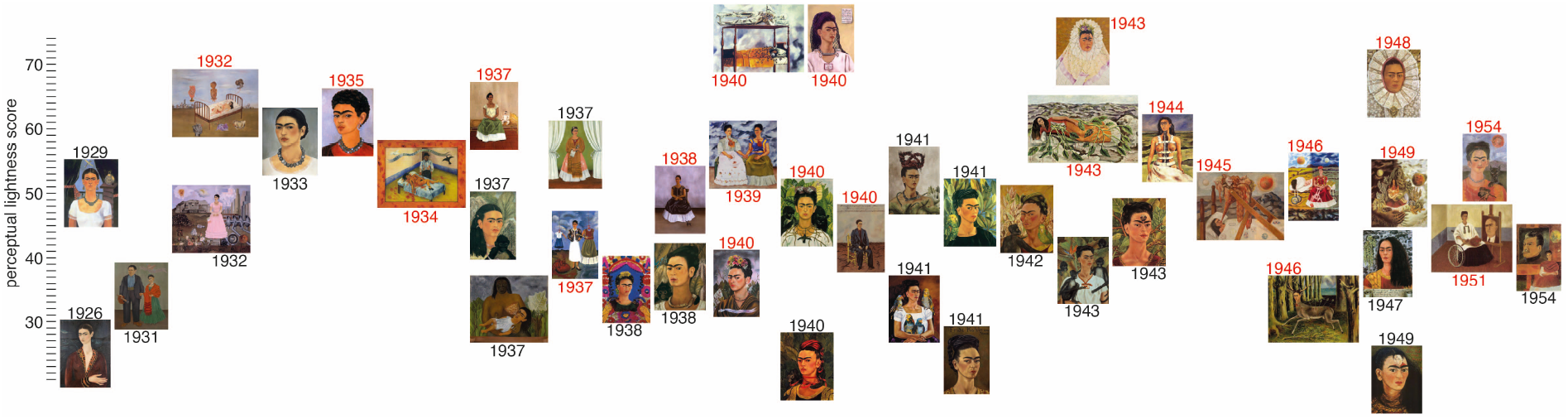
Timeline of the self-portraits of Frida Kahlo where the y-value of each canvas indicates the perceived luminance L*. The dates of the portraits that were scored 1 for physical or emotional pain are shown in red.

**Figure 2:**
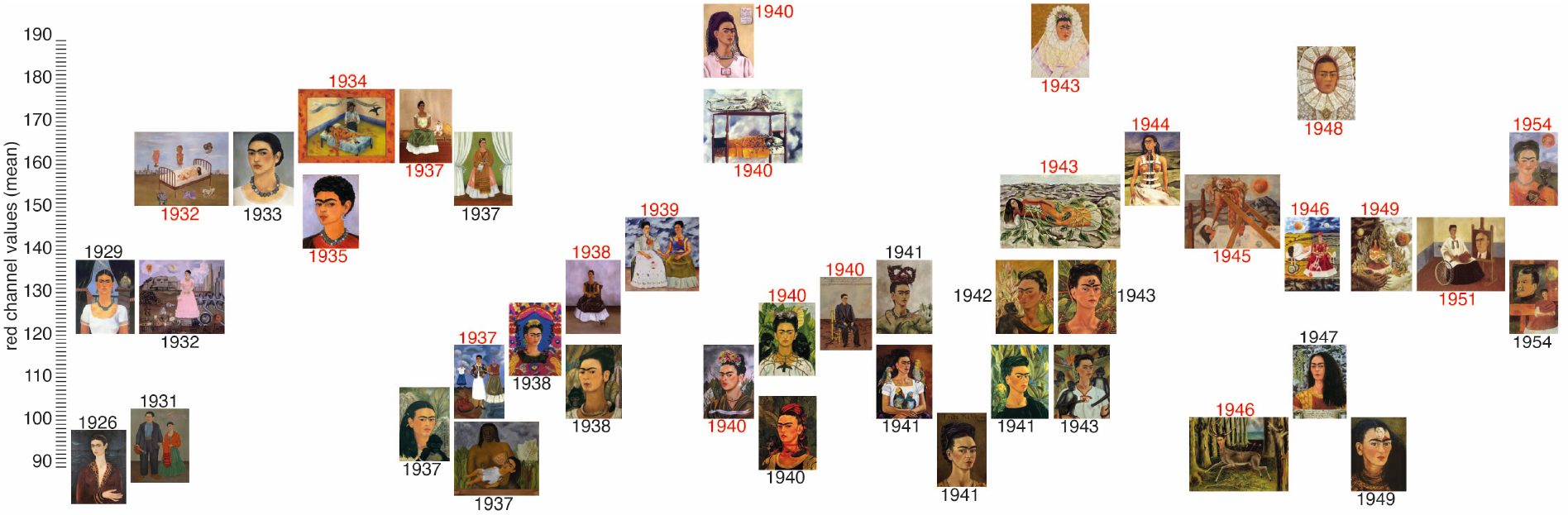
Timeline of the self-portraits of Frida Kahlo where the y-value of each canvas indicates the mean of the red channel R. The dates of the portraits that scored 1 for physical or emotional pain are shown in red.

**Figure 3:**
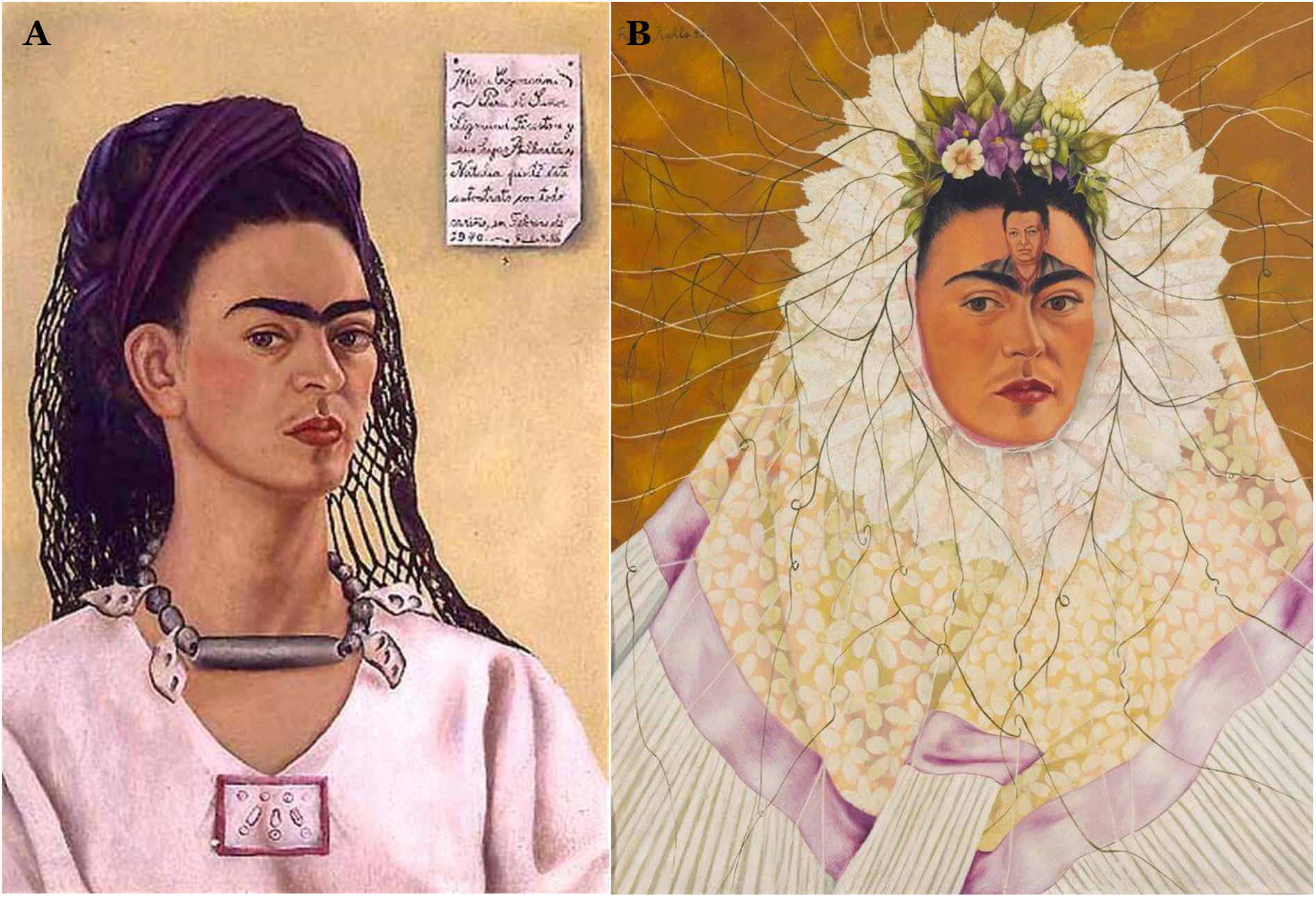
Two self-portraits of Frida Kahlo that scored the highest both in terms of redness and perceived luminance are A) “Self-portrait dedicated to Sigmund Firestone” (1940) and B) “Self-portrait as a Tehuana” (1943). Both were painted in 1940 (although the second one was completed in 1943) when, deeply hurt by the string of infidelities of her husband Diego Rivera, she filed for divorce. Frida went on to remarry him in December 1940 following the mutual agreement that they would not have sexual relations with each other, hence the image of the husband on the forehead indicating the intellectual obsession with Diego now contrasting with the physical detachment indicated by the nun-like costume ([15] pp.554, pp.567)

**Figure 4:**
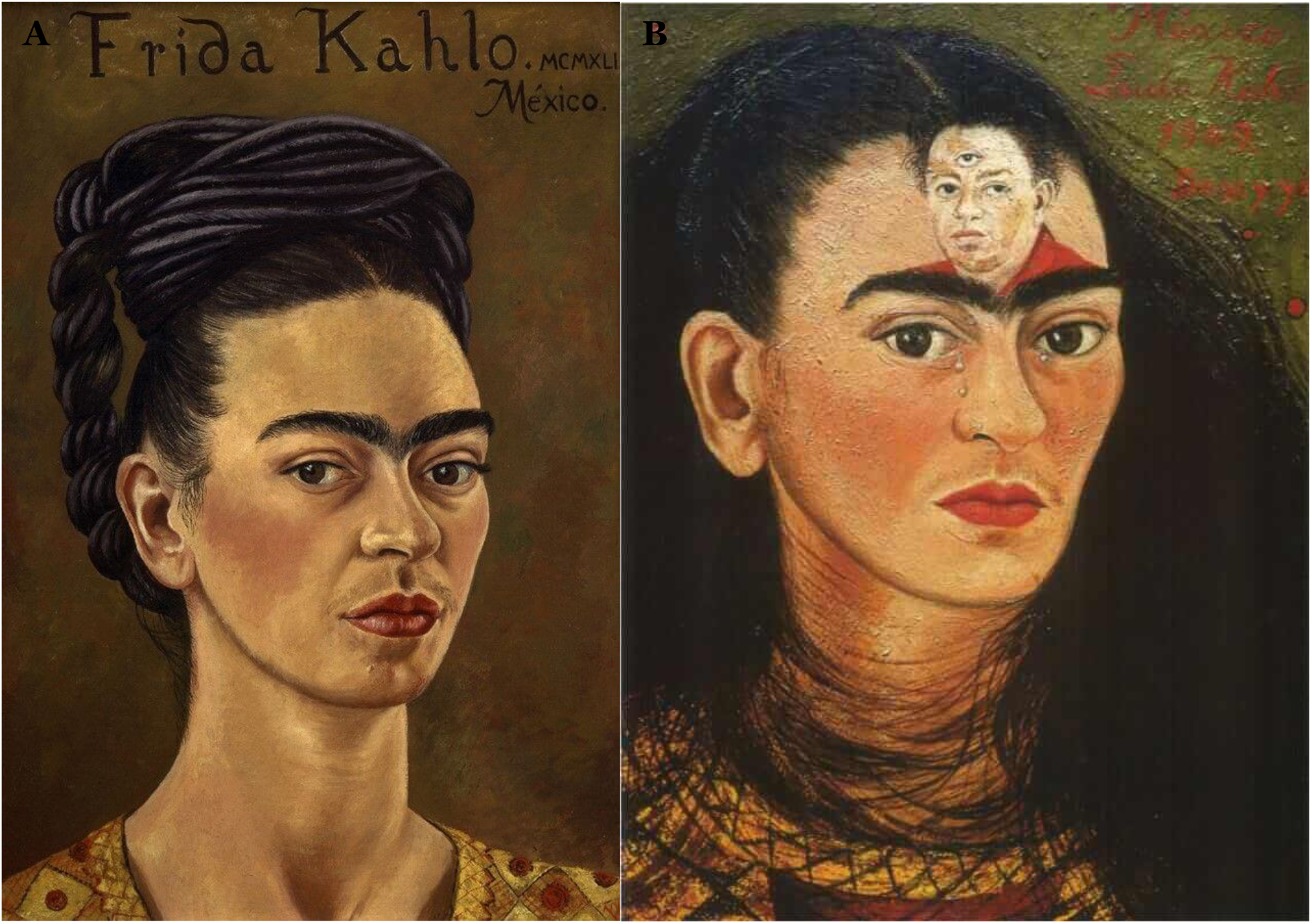
These two self-portraits have very similar themes to the ones shown in Figure 3 but they are at the bottom of both luminance and redness scales. They are A) “Self-portrait in a red and gold dress” (1941) and B) “Diego and I” (1949). They both were painted in times that were quieter with no noted physical distress although clearly the atmosphere is gloomy and sad (note the tears in B) [15].

The FD was clearly un-differentiated between the two groups. The timeline of the FD is shown in Figure 5 and, although it does not show a significant trend with time (i.e. correlation with years was not significant) it does however give insight into her style. Figure 6 illustrates the two outlying portraits, “Self-portrait with curly-hair” (1935), the one with the lowest FD, and “The broken column” (1944) that has the highest FD. The latter is of particular interest because it is commonly recognized as her masterpiece or, at least, the apex of her artistic journey that would afterwards take a painful turn because of the many surgical procedures she underwent ([15], p511). Indeed, the FD of the subsequent portraits show a substantial drop and recovers only before her sudden death in 1954. Finally, when the physical pain condition was contrasted with anger, no features were significantly different in the portraits.

**Figure 5:**
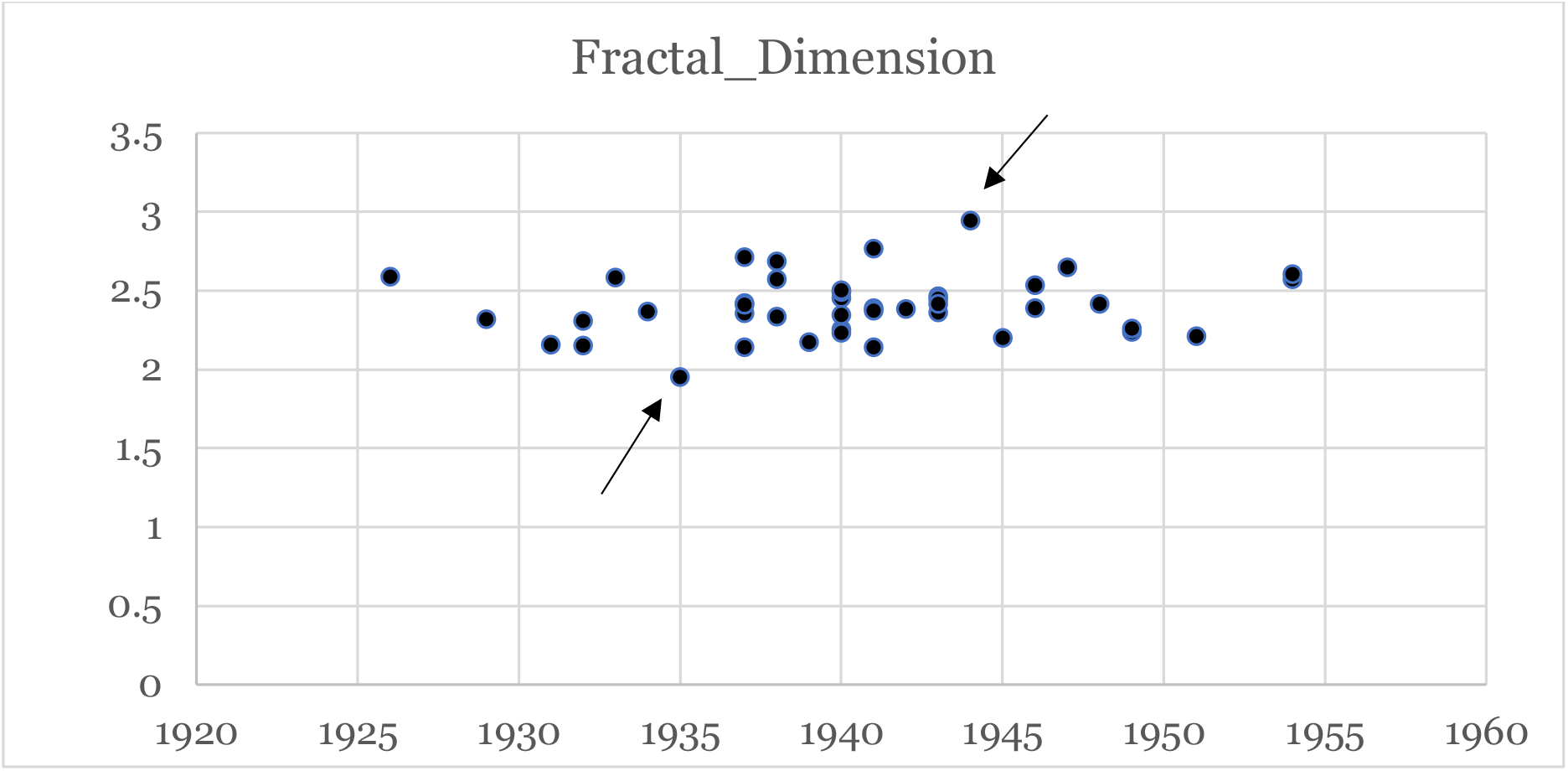
Plot of the fractal dimension FD across time. The data shows no trend and describes quantitatively the varying complexity of Frida’s work that remain constant, on average, along her professional life. There are extreme values indicated by arrows, one with value 1.9 produced in 1935 and one with FD=2.9 painted in 1944. These two portraits are shown in Figure 6.

**Figure 6:**
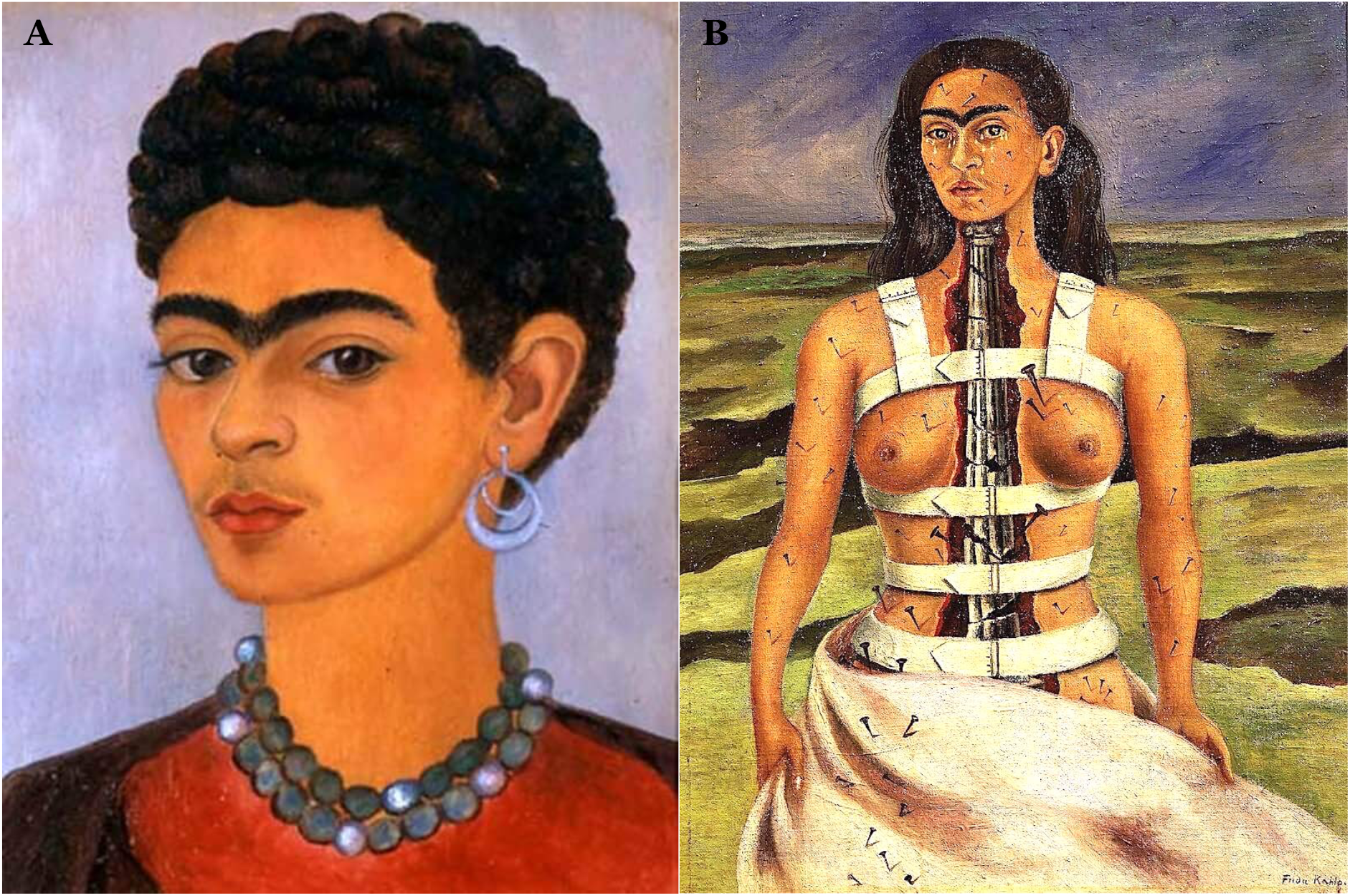
These two portraits A)” Self-portrait with curly-hair” (1935) and B) “The broken column” (1944) represent the two extremes in terms of canvas structuring in Frida Kahlo’s art. The first, inspired by Roman-Egyptian portraits, uses simple superposition of tones to convey the stern but dignified outrage of Frida towards the betrayal of her husband with her closest confidante, her sister Cristina. Note the cropped hair that was drawn in protest of Diego who liked her long hair ([15], p511). The second one, inspired by Greek architecture as by the ionic column standing for her damaged spine, is instead a highly structured piece. High FD quantifies the complexity of the color structure on canvas. This piece has been critically acclaimed for the realistic representation of her pain that runs across the whole representation from the rigidity of the spine that dominates the central part through the nails running down her leg; these depict the lumbar radiculopathy (or sciatica), a pain radiating down the leg along the dermatom. This piece is recognized as the climax of her art. ([15], pp.575).

## DISCUSSION

This work has used the personal and artistic timelines of Frida Kahlo to investigate the effects of physical and psychological distress on visual art.

The results, the first of their kind to our knowledge, confirm what has been suggested by experimental work, that there is an association between colours and physical and emotional pain [25; 33] The R channel was significantly increased in the pain/anger group of self-portraits but so were the G and B channels, the latter to a lower extent. In fact the data suggest that the association is driven not by hue but by the perceived lightness of the colours (whiteness) that, for the human eye, is higher for colours with low wavelength, red and yellow. This is further supported by it corresponding with an increase in luminance contrasts in the canvas.

The use of fractal analysis attempts a quantification of the impact of cognitive ailments on the artist’s artwork. The effect of pain on this marker was roughly constant across her lifetime, albeit with significant variability, which is likely to contain relevant information as the portrait that scored the highest FD value was her recognized masterpiece, “The broken column”. In our view, this indicates that, although pain was clearly reflected in Frida’s art, it did not compromise her technical or expressive ability.

In conclusion, this paper supports the value of quantitative image analysis in extracting psychological themes from visual art and in revealing the mental state of the artist. Hopefully these tools will become part of the clinical armoury of art intervention in in individuals with mental health problems cohorts in the future.

## Data Availability

All data produced in the present study are available upon reasonable request to the authors

http://www.FridaKahlo.org

## Acknowledgments

FET is supported by the National Institute for Health Research (NIHR) Biomedical Research Centre (BRC) at South London and Maudsley NHS Foundation Trust and King’s College London. All the authors declare to have no conflict of interest.

